# How to differentiate COVID-19 pneumonia from heart failure with computed tomography at initial medical contact during epidemic period

**DOI:** 10.1101/2020.03.04.20031047

**Authors:** Zhaowei Zhu, Jianjun Tang, Xiangping Chai, Zhenfei Fang, Qiming Liu, Xinqun Hu, Danyan Xu, Jia He, Liang Tang, Shi Tai, Yuzhi Wu, Shenghua Zhou

**Affiliations:** Department of Cardiovascular Medicine, the Second Xiangya Hospital, Central South University, Changsha, Hunan, China; Department of Emergency, the Second Xiangya Hospital, Central South University, Changsha, Hunan, China; Department of Radiology, the Second Xiangya Hospital, Central South University, Changsha, Hunan, China

**Keywords:** computed tomography, COVID-19, pneumonia, heart failure

## Abstract

**OBJECTIVES:** To compare chest CT findings in heart failure with those of Corona Virus Disease 2019 (COVID-19) pneumonia.

**BACKGROUND:** During epidemic period, chest computed tomography (CT) has been highly recommended for screening patients with suspected COVID-19. However, the comparison of CT imaging between heart failure and COVID-19 pneumonia has not been fully elucidated.

**METHODS:** Patients with heart failure (n=12), COVID-19 pneumonia (n=12) and one patient with both diseases were retrospectively enrolled. Clinical information and imaging of chest CT were collected and analyzed.

**RESULTS:** There was no difference of ground glass opacity (GGO), consolidation, crazy paving pattern, lobes affected and septal thickening between heart failure and COVID-19 pneumonia. However, less rounded morphology (8.3% vs. 67%, p=0.003), more peribronchovascular thickening (75% vs. 33%, p=0.041) and fissural thickening (33% vs. 0%, p=0.028), less peripheral distribution (33% vs. 92%, p=0.003) were found in heart failure group than that in COVID-19 group. Importantly, there were also more patients with upper pulmonary vein enlargement (75% vs. 8.3%, p=0.001), subpleural effusion and cardiac enlargement in heart failure group than that in COVID-19 group (50% vs. 0%, p=0.005, separately). Besides, more fibrous lesions were found in COVID-19 group although there was no statistical difference (25% vs. 0%, P=0.064)

**CONCLUSIONS:** Although there are some overlaps of CT imaging between heart failure and COVID-19, CT is still a useful tool in differentiating COVID-19 pneumonia.

---

In December 2019, an outbreak of Corona Virus Disease 2019 (COVID-19) caused by a novel coronavirus (severe acute respiratory syndrome coronavirus 2, SARS-CoV-2) began in Wuhan (Hubei, China) and spread rapidly(1). Recently, SARS-CoV-2 outbreaks in several other countries in the world. It seems a global pandemic is inevitable. Without efficient medicine, early detection and isolation becomes essential against novel coronavirus. However, as more secondary and tertiary cases have appeared, contact history and clinical manifestation may be not clear under certain circumstances. Chest computed tomography (CT) has been highly recommended for screening patients with suspected SARS-CoV-2(2).

Heart failure is the most common disease faced by emergency doctors and cardiologists. Pulmonary edema caused by heart failure can be appeared as exudative disease in CT scanning, which is, sometimes, difficult to be distinguished with other exudative disease in clinical practice. During epidemic period, it will be a great challenge for doctors to identify COVID-19 pneumonia from heart failure patients at the initial contact. So it is essential for cardiologists to be familiar with the imaging features of heart failure and COVID-19 pneumonia. Here, with the aim to enhance the diagnosis of COVID-19 pneumonia at initial medical contact, we present the clinical and imagine features of heart failure and COVID-19 pneumonia patients and one patient combined with these 2 diseases, to clarify the characteristics of heart failure and COVID-19 pneumonia.

## METHODS

### STUDY DESIGN AND PARTICIPANTS

This is a retrospective clinical study. We included consecutive patients with diagnosis of COVID-19 pneumonia or heart failure who had CT imaging at the initial medical contact at the Second Xiangya Hospital of Central South University from Dec. 1 to Feb. 28, 2020. Patients with negative CT findings and less than 18-years old were excluded. Available clinical history, laboratory, and epidemic characteristics were collected from 12 patients with heart failure and 12 patients with COVID-19 pneumonia. Besides, one patient with both heart failure and COVID-19 pneumonia was also enrolled. All the COVID-19 patients were diagnosed in fever clinics and then transferred to the designated hospital in Changsha, China.

The diagnosis and classification for heart failure is based on 2016 ESC Guideline(3) and the diagnosis of COVID-19 pneumonia was confirmed by the detection of SARS-CoV-2 in respiratory specimens with at least 3 positive results of qRT-PCR. Impaired heart function was defined as grade II or more in Killip or NYHA classification.

Clinical records, laboratory findings, and chest CT scans for all patients were obtained at initial medical contact. Two study investigators independently checked all the data. Throat-swab specimens from the upper respiratory tract were obtained. Maternal throat swab samples were collected and tested for SARS-CoV-2 with the Chinese Center for Disease Control and Prevention (CDC) recommended Kit (BioGerm, Shanghai, China), following WHO guidelines for qRT-PCR. qRT-PCR was repeated at least 3 times with different specimens.

This study was reviewed and approved by the Medical Ethical Committee of the Second Xiangya Hospital of Central South University (approval number 2020005), which waived the requirement for patients’ informed consent referring to the CIOMS guideline.

### IMAGINE INTERPRETATION

Two thoracic radiologists blinded to the clinical data reviewed the CT images independently and resolved discrepancies by consensus. All images were viewed on both lung (width, 1500 HU; level, −700 HU) and mediastinal (width, 350 HU; level, 40 HU) settings. The presence or absence of image features was recorded: ground-glass opacities (GGO), consolidation, rounded morphology, crazy paving pattern, peribronchovascular thickening, septal thickening, fissural thickening, small pulmonary vein enlargement, cardiac enlargement, lesion distribution, Lobes Involvement and bilateral lung disease. The detailed definitions of the above features were as described previously(4,5).

### STATISTICAL ANALYSIS

Statistical analysis was done with SPSS, version 25.0. Continuous variables were directly expressed as median, and interquartile range (IQR) values. Categorical variables were expressed as number (%). Means for continuous variables were compared using independent group t tests when the data were normally distributed; otherwise, the Mann-Whitney test was used. Proportions for categorical variables were compared using the χ2 test or Fisher exact test.

## RESULTS

### CLINICAL INFORMATION

The study included 12 patients with heart failure and 12 patients with COVID-19 pneumonia. The median age in patients with heart failure was 65 (IQR, 57,78) years and 5 (71%) were man. while the median age in patients with COVID-19 was 52 (IQR, 37,73) years and 8 (67%) were man. As shown in table 1. There was no difference in age, gender, oxygen saturation (SPO2) between two groups. Every patient in COVID-19 group had a contact history while no one had in heart failure group. For the symptoms, more patients got fever or respiratory symptoms in COVID-19 group than that in heart failure group (100% vs. 42%, p=0.005). Obviously, more patients had cardiovascular disease in heart failure group than that in COVID-19 group (100% vs. 17%, p<0.001). There were 6 patients with acute myocardial infarction, 4 patients with valve heart disease and 2 patients with cardiomyopathy in heart failure group. Consequently, higher NT-proBNP level was found in heart failure compared with that in COVID-19 group [119 (18.3, 1905) vs. 189 (64, 369), p=0.014]. Although there were no statistical differences of creatine kinase (CK) and CK-MB level, cardiac troponin T (cTnT) level in heart failure group was significantly higher than that in COVID-19 group [1793 (979,5852) vs. 0.06 (0.01, 0.13), p<0.001]. All the patients with heart failure were treated with anti-heart failure medicine in hospital. White blood cell and neutrophil count was higher in heart failure group compared with that in COVID-19 group [7.32(6.34, 9.91) vs. 5.1(4.3, 5.8), p<0.001; 5.46(4.35, 6.86) vs. 3.53(2.81,4.32), p=0.001]. Although lymphocyte count was decreased in both groups compared with the normal range, there was no statistical difference between groups. Besides, there was no statistical differences between groups in the level of inflammation markers including erythrocyte sedimentation rate (ESR), procalcitonin (PCT) and C-reactive protein (CRP). All the COVID-19 patients were confirmed by at least 3 times positive results of qRT-PCR for SARS-CoV-2, while the results of qRT-PCR for all the patients with heart failure were negative.

**Table 1.** Basic clinical information in patients with heart failure and COVID-19.

|  | **Heart failure** | **COVID-19** | ***P-value*** |
| --- | --- | --- | --- |
| **Age, years** | 65 (57,78) | 52(37, 73) | 0.126 |
| **Male** | 8 (67) | 8 (67) | 1.0 |
| **Epidemic contact** | 2 (17) | 12 (100) | <0.001 |
| **Fever or respiratory symptoms** | 5 (42) | 12 (100) | 0.005 |
| **SPO2(%)** | 94±2 | 93±3 | 0.08 |
| **Cardiovascular disease** | 12 (100) | 2(17) | <0.001 |
| Acute myocardial infarction | 6 (50) | 0 |  |
| Valve heart disease | 4 (33) | 2 (17) |  |
| Cardiomyopathy | 2 (17) | 0 |  |
| **Impaired heart function** | 12(100) | 3 (25) | <0.001 |
| **II** | 5 | 3 |  |
| **III** | 5 | 0 |  |
| **IV** | 2 | 0 |  |
| **While blood cell count (×10<sup>9</sup>)** | 7.32(6.34, 9.91) | 5.1(4.3, 5.8) | <0.001 |
| **Neutrophil count (×10<sup>9</sup>)** | 5.46(4.35, 6.86) | 3.53(2.81,4.32) | 0.001 |
| **Lymphocyte count (×10<sup>9</sup>)** | 1.03 (0.78, 1.55) | 0.83 (0.68,1.14) | 0.196 |
| **CRP (mg/l)** | 15.6 (4.9, 105.5) | 26.7 (12.8, 41.2) | 0.951 |
| **ESR (mm/h)** | 47 (29.5, 92.3) | 23 (5.3, 56) | 0.394 |
| **PCT (ng/ml)** | 0.094 (0.039, 0.473) | 0.085 (0.045, 0.101) | 0.770 |
| **CK (u/l)** | 78.7 (27.8, 325) | 91.3 (57.6,121) | 0.622 |
| **CK-MB (u/l)** | 15.9 (11.2, 22.4) | 10.5 (7.2,13.4) | 0.056 |
| **cTnT (pg/ml)** | 119 (18.3, 1905) | 0.06 (0.01,0.13) | <0.001 |
| **NT-proBNP(pg/ml)** | 1793 (979,5852) | 189 (64,369) | 0.014 |
| **SARS-CoV-2 positive** | 0 (0) | 12 (100) | <0.001 |

Values are median (interquartile range), n (%), or mean ± SD. SPO2= oxygen saturation, CRP= C-reactive protein, ESR= erythrocyte sedimentation rate, PCT= procalcitonin, CK= creatine kinase, CK-MB= creatine kinase-MB, cTnT= cardiac troponin T.

### IMAGING FEATURES

Figure 1 showed the typical imaging for heart failure (figure 1A, 1B) and COVID-19 pneumonia (figure 1C, 1D). However, there were some similarities and differences between these 2 groups. The comparison of imaging features was concluded in Table 2. Compared with patients with COVID-19, 11 patients in 12 had GGO in heart failure group, and there was no statistical difference between two groups (92% vs. 83%, p=0.537). There were also no differences between groups for consolidation, crazy paving pattern, and septal thickening. Hoverer, less patients had rounded morphology (8.3% vs. 67%, p=0.003) (figure 1C, 1D), more patients had peribronchovascular thickening (75% vs. 33%, p=0.041) (figure 2A, 2B) and fissural thickening (33% vs. 0%, p=0.028) (figure 1B, 2A) in heart failure group than that in COVID-19 group. For lesion distribution, although there was no difference of central distribution between groups, patients with heart failure were less like to have a peripheral distribution (33% vs. 92%, p=0.003). Importantly, 6 patients in 12 had gravity dependent gradient distribution in heart failure group (figure 1A), while no patient had such distribution in COVID-19 group (p<0.001). For the lobes affected, there was no statistical difference in the frequency of 2 or more lobes involvement and bilateral lung disease. Importantly, there were more patients with pulmonary vein enlargement in heart failure group compared with that in COVID-19 group (75% vs. 8.3%, p=0.001) (figure 2C). And there were more patients with subpleural effusion and cardiac enlargement in heart failure group than that in COVID-19 group (50% vs. 0%, P=0.005, separately). Besides, more fibrous lesions were found in COVID-19 group although there was no statistical difference (25% vs. 0%, P=0.064) (figure 2E). Figure 2 showed different imaging features for heart failure (2A, 2B, 2C) and COVID-19 (2D, 2E, 2F).

**Table 2.** Comparison of imaging features of heart failure and COVID-19 pneumonia.

|  | **Heart failure** | **COVID-19** | ***P-value*** |
| --- | --- | --- | --- |
| Ground-Glass Opacities | 11(92) | 10(83) | 0.537 |
| Rounded morphology | 1(8.3) | 8(67) | 0.003 |
| Fibrous lesions | 0(0) | 3(25) | 0.064 |
| Consolidation | 6(50) | 8(67) | 0.408 |
| Crazy Paving Pattern | 3(25) | 4(33) | 0.653 |
| Pulmonary vein enlargement | 9(75) | 1(8.3) | 0.001 |
| Peripheral Distribution | 4(33) | 11(92) | 0.003 |
| Gradient distribution | 6(50) | 0(0) | <0.001 |
| Central Distribution | 4(33) | 2(17) | 0.346 |
| Septal thickening | 11(92) | 9(75) | 0.273 |
| Fissural thickening | 4(33) | 0(0) | 0.028 |
| Peribronchovascular thickening | 9(75) | 4(33) | 0.041 |
| Subpleural effusion | 6(50) | 0(0) | 0.005 |
| Cardiac enlargement | 6(50) | 0(0) | 0.005 |
| More than 2 lobes affected | 8(67) | 6(50) | 0.408 |
| Bilateral Lung Disease | 10(83) | 6(50) | 0.083 |
| 2 or more Lobes Involvement | 11(92) | 8(67) | 0.132 |
| Right upper lobe | 6(50) | 5(42) | 0.682 |
| Right middle lobe | 8(67) | 4(33) | 0.102 |
| Right lower lobe | 10(83) | 7(58) | 0.178 |
| Left upper lobe | 9(75) | 8(67) | 0.653 |
| Left lower lobe | 9(75) | 8(67) | 0.653 |

Values are n (%).

**FIGURE 1.**
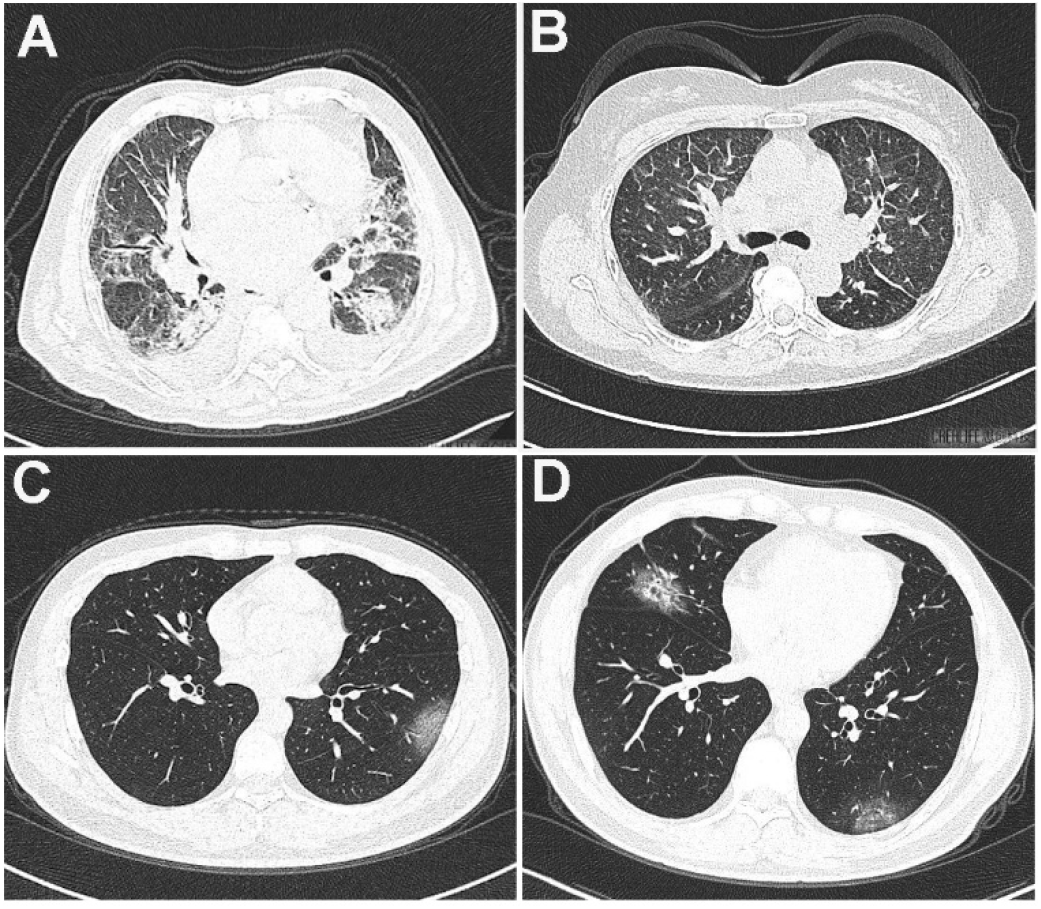
Typical imaging for heart failure and COVID-19 pneumonia. A, Heart failure with bilateral diffuse disease. The disease presented with a gradient distribution and partial GGO and consolidation inside, accompanied with subpleural effusion. Peribronchovascular and septal thickening were also found. B, Heart failure with bilateral diffuse GGO disease. Peribronchovascular thickening, clear interlobular septal thickening even fissural thickening were found without subpleural effusion. C, COVID-19 with single rounded subpleural GGO in left lung. D, COVID-19 with rounded GGO in bilateral lungs, partial consolidation was found inside.

**FIGURE 2.**
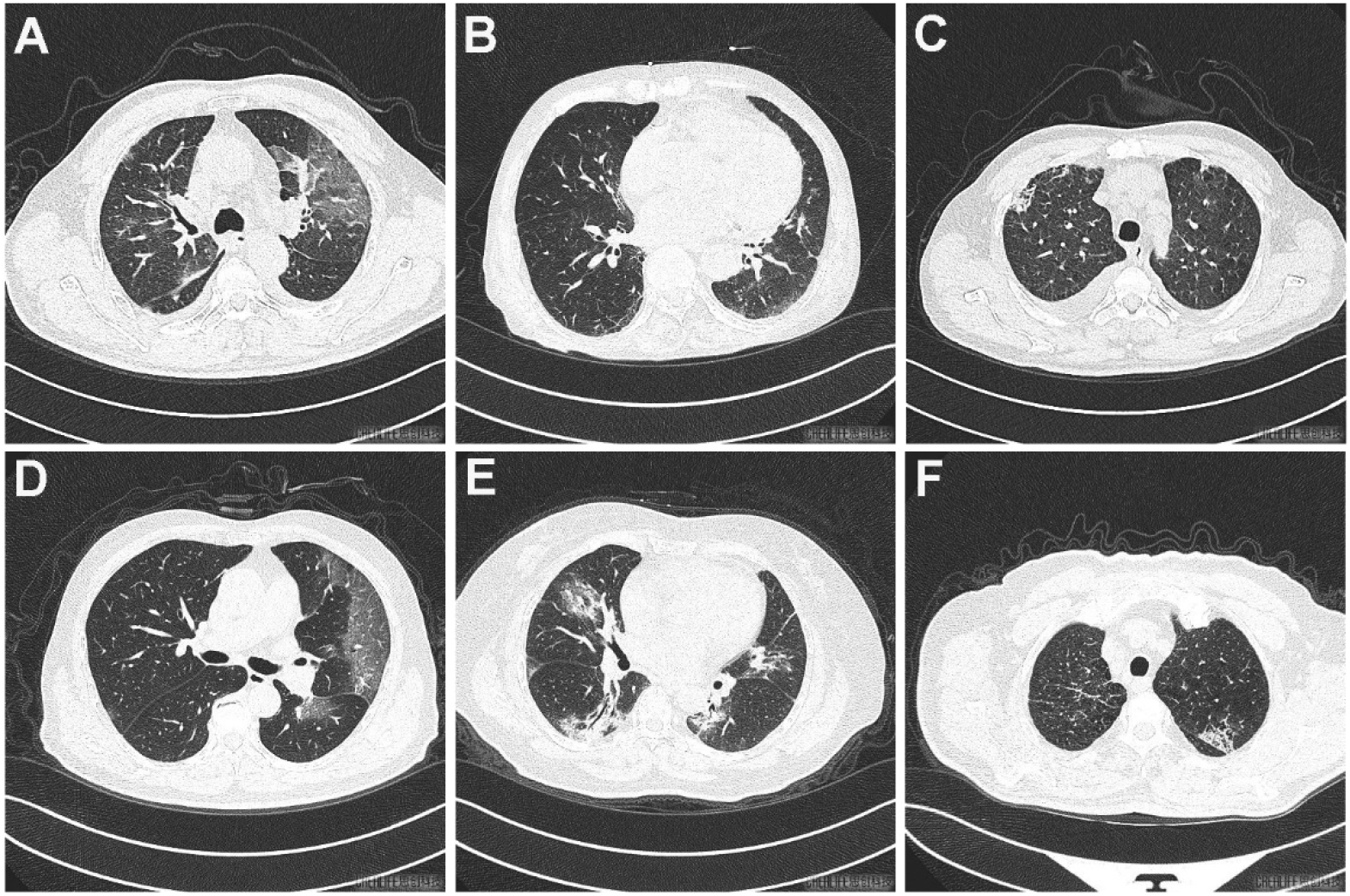
Different imaging features for heart failure and COVID-19. A, Big patchy GGO in left lung and sporadic GGO in right lung. Peribronchovascular thickening, fissural thickening (arrow) were also found with subpleural effusion. B, Subpleural GGO with band-shaped morphology (arrow) in the dorsal segment of bilateral lungs. Interlobular septal thickening and cardiac enlargement were also found. C, Patchy lesion of Crazy Paving Pattern in right upper lung with partial consolidation inside. Small pulmonary vein enlargement (arrow) was also found, accompanied with subpleural effusion in the right lung. D, Big patchy GGO in left lung. Interlobular septal thickening was also found inside. E, Multiple disease mixed with GGO and consolidation in bilateral lungs. Fibrous lesion (arrow) was found in the left lung. F, Patchy lesion of Crazy Paving Pattern in left upper lung with partial consolidation inside.

### A CASE WITH BOTH HEART FAILURE AND COVID-19 PNEUMONIA

A 64-year-old male with recent travel history to Hubei, China, the epicenter of the SARS-CoV-2 outbreak, was admitted to the hospital with fever and orthopnea. He was diagnosed with coronary artery disease in last year and was stented in Left anterior descending artery. The chest CT (Figure 3) showed multi-focal GGO with parenchyma consolidation and subpleural effusion, predominantly involving upper lungs. There was no rounded morphology, while the Septal and bronchial wall were thickened. The third swap test of SARS-CoV-2 was positive 1 week after CT scanning.

**FIGURE 3.**
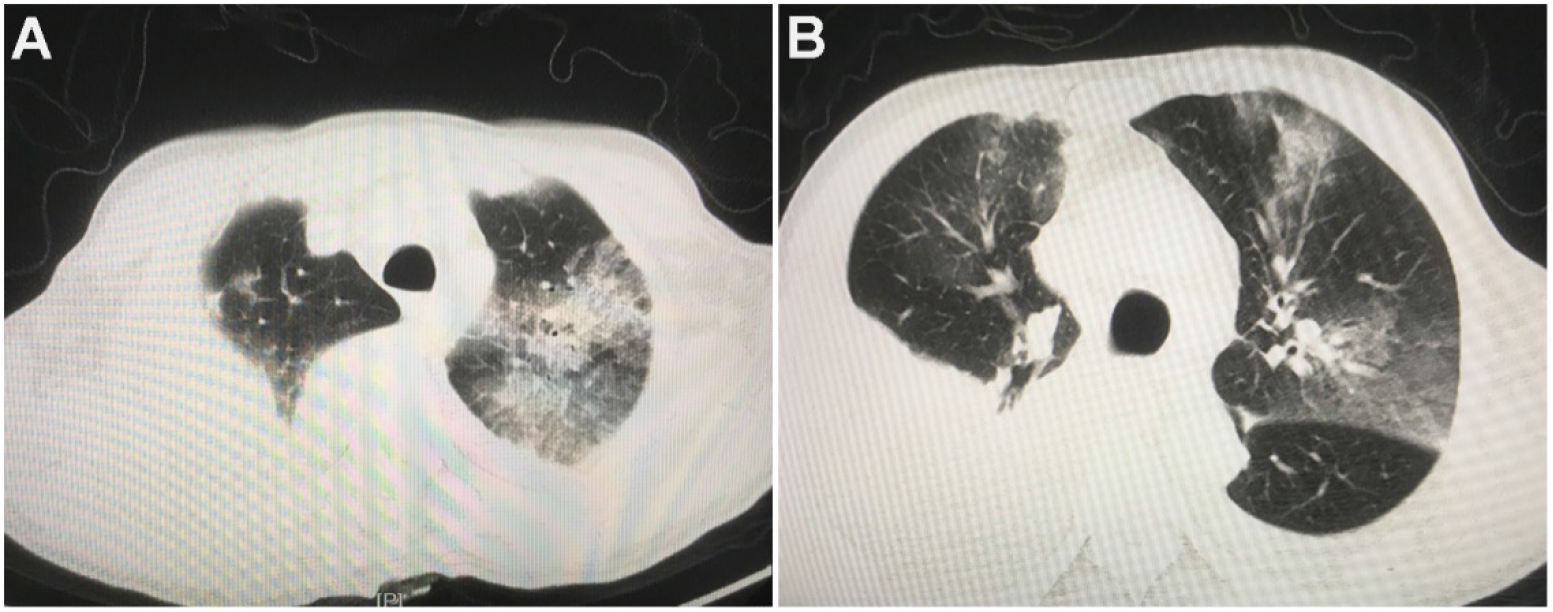
CT imaging of a patient with both heart failure and COVID-19. A, Diffuse disease mixed with GGO and consolidation in bilateral upper lungs, accompanied with interlobular septal thickening and subpleural effusion. B, Big patchy GGO with irregular morphology in bilateral lungs. Peribronchovascular thickening and subpleural effusion were also found.

## DISCUSSION

In this study, under a special situation of COVID-19, the clinical characteristics of heart failure and COVID-19 pneumonia were concluded, and the similarity and differences of CT imaging features were also verified. We found that GGO and septal thickening were all common lesions in both diseases. Significant differences exist in the distribution type, lesion morphology, upper small pulmonary vein enlargement, fissural thickening, peribronchovascular thickening, subpleural effusion and cardiac enlargement.

It is essential to identify the suspected patients as early as possible to control the spread of the disease. Up to present, China has updated to the 7^th^ edition of the diagnosis for suspected patients. Contact history, clinical manifestation (fever or respiratory symptoms), lab test (white blood cell count normal or decreased, lymphocyte count decreased) and imagine of pulmonary CT are included in the evaluation. A patient can be suspected if fulfill contact history and any 2 of the latter 3. Even for patients without clear contact history, the latter 3 can be as evidence for suspected patients. For patients with heart failure in the absence of fever, chest uncomfortable or chest pain or apnea sometimes combined with respiratory symptom. In this study, 4 patients in 12 have respiratory symptoms. Besides, nearly half of COVID-19 patients may not have fever at admission(6) and the lymphocyte count in patients with heart failure can also be decreased. So pulmonary CT as an objective method will be a key point for patients without clear contact history and typical symptoms.

In this study, we found that there are some similarities in imaging features for patients with heart failure and COVID-19 pneumonia. Both diseases had GGO, consolidation, crazy paving pattern and septal thickening, which will make it difficult to differentiate heart failure from COVID-19 by CT scanning. However, with careful checking of the imaging, the distribution type of the two kinds of diseases were significantly different. Heart failure were more likely to have a central and gravity associated gradient distribution, while COVID-19 usually have more peripheral distribution. Importantly, there are more lesions with rounded morphology in COVID-19 than that in heart failure. In detail, although both diseases have septal thickening, heart failure usually has more peribronchovascular thickening and interlobular septal even fissural thickening. While COVID-19 usually affected the smaller septum. This may be attributed to the different severity of the 2 disease at the beginning, which means heart failure may progress more rapidly at first. COVID-19 usually progresses with different imaging features at different stages last for about 2-3 weeks(7). At first GGO is the predominant feature and gradually spread and consolidated, at last stage, the consolidation will be absorbed. This study was just focus on the initial medical contact, and the CT imaging showed here were almost belong to the early stage. So predominated GGO and some consolidation were most popular in COVID-19. Although fibrous lesions rare reported by now, 3 cases with COVID-19 were found with such lesions in this study, which is still consistent with current research(8). Interestingly, the CT imaging characteristics of both diseases may be mixed in one patient with heart failure and COVID-19, or the features of COVID-19 at initial medical contact will be covered by more progressively heart failure. So it should be more careful to contact with a patient with heart failure in epidemic area.

Hydrostatic pulmonary edema manifested by interstitial edema and alveolar flooding is the most frequently recognized pathophysiologic and radiologic feature in heart failure(9). These features are virtually identical for left-sided heart failure and fluid overload. Interstitial edema occurs first with an increase of 15-25mm hg in mean transmural arterial pressure and results in the early loss of definition of subsegmental and segmental vessels, mild enlargement of the peribronchovascular spaces, and subpleural effusions. With pressure increasing, alveolar flooding will occur. The pathophysiology of central distribution can be explained by follows: increased tissue hydration which allows water to easily flow centrally, the pumping effect of the respiratory cycle which causes overall fluid flow toward the hilum, contractile property of alveolar septa allows them to expel interstitial edema toward the hilum. Diffuse bilateral interstitial and/or interstitial-alveolar infiltrates are most commonly caused by viruses. Different with heart failure, COVID-19 was characterized by an inflammatory pulmonary edema and alveolar damage. Recently, fibromyxoid exudation has been found in the lung tissue from patient obtained at autopsy(10), which is absent in heart failure. All those above make the imaging differences from heart failure.

In this study, the leukocyte and neutrophil count of patients with heart failure was significantly higher than that of covid-19 patients, which may be attributed to the inflammatory state in vivo induced by heart failure or acute myocardial infarction(11,12). In addition, other inflammatory indexes such as erythrocyte sedimentation rate and C-reactive protein in both groups also increased compared with the normal range. The results are consistent with previous studies(13,14). Interestingly, the lymphocyte count was also decreased in heart failure, which may because, on one hand, the increase of neutrophil ratio in patients with heart failure may cause the relative decrease of lymphocyte ratio. On the other hand, it may be related to the increase of cortical hormone level after stress response.

There are several limitations of this study. First, this is a retrospective study with limited cases. Most of the patients with COVID-19 enrolled were non-severe cases. Secondly, we mainly focus on the clinical and imaging features at the initial medical contact, while the dynamic changes with appropriate therapy are definitely helpful to differentiate the 2 diseases.

In conclusion, during epidemic period, it is essential for doctors to identify the imaging features of COVID-19 and heart failure. Although both diseases can have similar GGO and septal thickening, rounded morphology, peripheral distribution and fibrous lesion were relatively specific in COVID-19. While heart failure usually has more peribronchovascular thickening, fissural thickening, subpleural effusion and cardiac enlargement.

## Data Availability

N/A

